# Elucidating the Dynamics and Impact of the Gut Microbiome on Maternal Nutritional Status During Pregnancy in Rural Pakistan: Study Protocol for a Prospective, Longitudinal Observational Study

**DOI:** 10.1101/2023.09.11.23295048

**Authors:** Yaqub Wasan, Jo-Anna B. Baxter, Carolyn Spiegel-Feld, Kehkashan Begum, Arjumand Rizvi, Junaid Iqbal, Jessie M. Hulst, Robert Bandsma, Shazeen Suleman, Sajid B. Soofi, John Parkinson, Zulfiqar A. Bhutta

## Abstract

**Introduction:** Undernutrition during pregnancy is linked to adverse pregnancy and birth outcomes and has downstream effects on the growth and development of children. The gut microbiome has a profound influence on the nutritional status of the host. This phenomenon is understudied in settings with a high prevalence of undernutrition, and further investigation is warranted to better understand such interactions.

**Methods:** This is a prospective, longitudinal observational study to investigate the relationship between prokaryotic and eukaryotic microbes in the gut and their association with maternal BMI, gestational weight gain, and birth and infant outcomes among young mothers (17-24 years) in Matiari District, Pakistan. We aim to enroll 400 pregnant women with low and normal BMIs at the time of recruitment (<16 weeks of gestation).

**Analysis:** To determine the weight gain during pregnancy, maternal weight is measured in the first and third trimesters. Gut microbiome dynamics (bacterial and eukaryotic) will be assessed using 16S and 18S rDNA surveys applied to the maternal stool samples. Birth outcomes include birthweight, SGA, LGA, preterm birth, and mortality. Infant growth and nutritional parameters include WHO z-scores for weight, length, and head circumference at birth through infancy. To determine the impact of the maternal microbiome, including exposure to pathogens and parasites on the development of the infant microbiome, we will analyze maternal and infant microbiome composition, micronutrients in serum using metallomics (e.g., zinc, magnesium, and selenium), and macronutrients in the stool. Metatranscriptomics metabolomics and markers of inflammation will be selectively deployed on stool samples to see the variations in dietary intake and maternal nutritional status. We will also use animal models to explore the bacterial and eukaryotic components of the microbiome.

**Ethics and dissemination:** The study is approved by national and institutional ethics boards, and findings will be published in peer-reviewed journals.

**Study registration:** ClinicalTrials.gov Identifier: NCT05108675.

**Strengths and limitations:**

- The study targets the high fertility age group (17-24) with almost half cohort consisting of low BMI mothers, potentially with an additional risk of adverse pregnancy outcomes, providing an opportunity to comprehend the systematic understanding of the role of microbiota in several pregnancy, birth, and infant outcomes.
- Study investigates both prokaryotic and eukaryotic dynamics of the gut microbiome for in-depth mechanistic insights in a highly malnourished population where contextual evidence is rare.
- Longitudinal design and data collection on a range of exposure indicators and biochemical analysis would enable us to evaluate the association of gut dynamics with several physiological and environmental factors.
- The study follows the STROBE guidelines; however, we expect controlling for all confounding variables may not be possible.
- Focusing on young women, 17-24 years of age, the findings may not be generalizable to younger or older demographics.

## Introduction

Undernutrition during pregnancy is associated with an increased risk of poor birth outcomes and intra-uterine growth restriction of the fetus [1] and globally, is a leading cause of death in under five children [2] as well as maternal morbidity and mortality[3, 4]. Moreover, women with low body mass index (BMI) are at increased risk for adverse pregnancy and birth outcomes [5]. Undernutrition before or during pregnancy may also have a long-term impact on the offspring [6-8]. Among women in low and middle-income countries (LMICs), several micronutrient deficiencies often co-exist due to insufficient dietary intake [1]. Pre-existing micronutrient deficiencies may be exacerbated during pregnancy as a result of the increased metabolic requirements [9]. In particular, adolescent pregnancies are associated with a 50% increased risk of stillbirths and neonatal deaths, and an increased risk of preterm birth, low birth weight (LBW), and asphyxia [10-12]. In Pakistan, The National Nutritional Survey (2018) revealed majority of women of reproductive age had some form of micronutrient deficiency, for example, 42.7% were anemic. Nationwide, 40.2% of children under 5 were stunted, and 17.7% had severe wasting [13]. Within the current study area, a previous multicentre study showed stillbirths and neonatal mortality rates in district Matiari [14] were higher than the national rates [15].

The intestinal microbiome has emerged as a key factor affecting nutritional status, with impaired maturation contributing to undernutrition [16-22]. During pregnancy, dramatic changes to the gut microbiota occur, with a decrease in individual (alpha) diversity but an increase in population (beta) diversity [23]. Relative to the first trimester, microbiota in the third-trimester exhibit higher abundances of *Proteobacteria*, typically associated with obesity in humans, and when transplanted to mice, result in increased adiposity and insulin insensitivity. Such adaptations may increase energy extraction from the diet to support pregnancy [24], raising interest in dietary supplements to improve pregnancy outcomes [25].

Maternal environmental enteric dysfunction (EED) during pregnancy is shown to adversely impact birth outcomes [26]. Cumulative pathogen exposures in children confer a high risk for poor growth [27, 28] and EED [29-31]. EED is thought to be triggered by dysbiosis [32, 33], initiated by nutrient deficiencies, antibiotic treatment, and/or pathogen exposure. This may further exacerbate pathogen colonization, impair development of the mucosal immune system, and disrupt, by as yet unknown mechanisms, metabolic processes that supply nutrients and energy for normal growth [19]. Variability in microbial abundance and taxa in infancy and second year of life in association with exposure to different interventions and environment (rural and urban) in Pakistani children also warrant in depth analysis include metagenomics and metabolomics to identify microbiota directed interventions to address malnutrition. [34]

Thus, this study aims to systematically investigate how shifts in the gut microbial community impacts nutritional status during pregnancy in young women. Leveraging whole microbiome RNASeq (metatranscriptomics) and metabolomics, our overall aim is to further develop mechanistic insights into the relationships between host socio-environmental factors, nutritional status, and microbiome dynamics during pregnancy, and how they contribute to birth outcomes and infant growth during the first year of life. Study findings may help devise microbiota-informed solutions to improve nutritional status of mothers during pregnancy and improve birth outcomes and infant health, well-being, and survival.

### Objectives

The primary aim of this study is to assess if alterations of the microbiota in the maternal gut (dysbiosis) are associated with maternal gestational weight gain, and to determine the association between maternal microbiome dysbiosis during pregnancy and birth outcomes (i.e., birth weight, preterm births, small for gestational age, large for gestational age), infant growth, nutritional status, and health status in the first year of life. Several secondary outcomes are integrated to better understand the influence of maternal factors such as dietary intake, maternal BMI, and exposure to pathogens on the gut health and microbiome of infants. Finally, this study will explore how socio-economic factors; including gender, poverty, exclusion, and empowerment influence the health of a mother’s microbiome. Details are provided in supplementary material Table 1.

**Table 1:** Overview of study visits, visit activities, data, and specimen collection.

| Procedures | Pregnancy Surveillance | MV1: 10-14 Weeks Post-Conception | MV2: 30-34 Weeks Post-Conception | MV3: Maternal 24-72 Hours Post-Partum | IV1: Infant 24-72 Hours Post-delivery (same visit as of mom) | MV4: Maternal 3 months post-partum | IV2: Child 3-Months (same visit as of mom) | MV5: Maternal 6 Months Post-Partum | IV3: Infant 6 Months (same visit) | MV6: Maternal 12-Months Post-Partum | IV4: Child 12-Months (same visit as of mom) |
| --- | --- | --- | --- | --- | --- | --- | --- | --- | --- | --- | --- |
| Pregnancy Surveillance | X |  |  |  |  |  |  |  |  |  |  |
| Screening and Confirmation of Pregnancy |  | X |  |  |  |  |  |  |  |  |  |
| Written Informed Consent |  | X |  |  |  |  |  |  |  |  |  |
| Household and individual demographics |  | X |  |  |  |  |  |  |  |  |  |
| 24-hour food recall |  | X | X |  |  |  |  |  |  | X |  |
| Minimum Dietary Diversity Score for Women (MDD-W) |  | X | X |  |  | X |  |  |  | X |  |
| Infant feeding practices |  |  |  |  |  |  | X |  | X |  | X |
| Anthropometric Measurements |  | X | X | X | X | X | X | X | X | X | X |
| Empowerment |  | X |  |  |  | X |  |  |  |  |  |
| Birth and Labour History |  |  |  | X | X |  |  |  |  |  |  |
| Health and Medication Use Assessment |  |  | X | X | X | X | X | X | X | X | X |
| Stool Sample |  | X | X |  |  | X | X |  |  | X | X |
| Blood Sample |  | X | X |  |  |  |  |  |  | X | X |
| MV= Maternal visit; IV= Infant visit |  |  |  |  |  |  |  |  |  |  |  |

### Study hypothesis

The study hypothesizes that the alterations of the microbiota in the maternal gut (dysbiosis) exacerbated by nutritional status or pathogen exposure during pregnancy, impact weight gain during pregnancy because of impaired nutrient absorption, leading to corresponding negative birth outcomes.

## Methods

### Design

This study employs a prospective, longitudinal observational design to be able to capture maternal and infant outcomes over time.

### Study setting and population

The study is being conducted in a rural district, Matiari, within Sindh province in Pakistan. Matiari is 200 kilometers away from Karachi and includes more than 1400 villages and a population of about 800,000. The study setting has a well-established community and health system liaison, basic demographic surveillance, and field centers for research. This district is representative of typical conditions in Pakistan, and there is a close working relationship with the community, civic society leaders, and public health departments. The study aims to collect blood and stool samples from pregnancy to 1-year post-partum, to monitor the dynamic relationships between microbiome community structure and function with gut health and host nutritional status.

At the core of this study are two complementary cohorts of young women, one in Matiari, Pakistan and one in Toronto, Canada. Here we focus on the Pakistani cohort. We will recruit young, married women, including newlyweds, 17-24 years of age, living in Matiari District, Pakistan. We focused on this younger demographic due to our lack of knowledge on the microbiome of young women, and their increased vulnerability to undernutrition; two in every five young women (15-24) living in Matiari are underweight [34], many exhibit suboptimal dietary diversity [35], and more than 90% experience at least one form of micronutrient deficiency [36]. The cohort, based in Toronto is focused on immigrant and refugee populations and will be described elsewhere.

### Study status

The first participant was enrolled on November 25, 2021. Data collection for the study is expected to be completed in 2024.

### Sample size

We aim to recruit 400 young women 17-24 years of age into two groups based on their BMI at timing of pregnancy identification: group 1 will include those with a ‘normal ‘BMI (i.e., BMI between 18.5 to 25 kg/m^2^) and group 2 will include those with an underweight BMI (i.e., <18.5 kg/m^2^), as per World Health Organization’s (WHO) guideline definitions [37]. The adequacy of the sample size was verified using the ‘pwr’ package (version 1.2-2) in R (version 3.6.1). Calculations were based on the correlation between *α*-diversity (Shannon index) and weight gain during pregnancy. Assuming 400 participants are recruited into the study, and a type I error rate of 0.05, there will be 80% power to detect a correlation coefficient (*r*) >0.14. This is conventionally considered a small effect size [38]. Thus, we expect to be powered to reveal a significant association between weight gain during pregnancy and microbial diversity.

### Identification of pregnancies

To identify pregnancies within the study area, we leveraged the network that we had established in a previous trial, the Matiari emPowerment and Preconception Supplementation (MaPPS) Trial [39, 40]. Using existing participant lists, women and families are initially approached by phone to get information on early pregnancies. Volunteers from within local villages and primary health care facilities area are also encouraged to report new pregnancies. Verbal consent is taken from the identified pregnant women to share their pregnancy status with the study team for further eligibility screening. Research field staff also perform random checks of households and villages and meet with lady health workers and volunteers in the field to get information on pregnancies.

### Eligibility screening

Pregnancy identification data is assembled at the field office on daily basis. The study team contacts the pregnant women for eligibility screening at their homes. The study staff obtains verbal consent to take the potential participant’s height and weight to determine the anthropometric eligibility criteria (BMI <25 kg/m^2^) and to conduct a pregnancy test to confirm pregnancy. Women 17-24 years of age who are in good general health, without known chronic diseases, and <16 weeks of gestation are invited to take part in the study. To participate, women are asked to provide written consent and agree to comply with the study procedure. Those with a BMI ≥25 kg/m^2^ at the time of recruitment and/or, participating in another nutritional trial, and/or report taking antibiotics in the last three months, and/or screen for potential signs of COVID-19 are not eligible for study participation. Only one participant per household is enrolled in the study, given the potential for within household dietary and microbial similarities.

### Participant recruitment

Pregnant women who meet the eligibility criteria are formally invited for study participation following written informed consent. Informed consent is administrated pursuing Aga Khan University’s (AKU) well-established protocols for research ethics compliance. Participants are also explained the option to sign an additional section of the consent form for future genetic testing of biospecimens. Participants are given the option to discuss the consent form with their families, before agreeing to participate in the study.

### Data and specimen collection

All data collection is completed by trained female data collectors at the participant’s home. This includes verbal data collection, anthropometric assessments, and blood and stool sample collection. Data collection at birth, which is collected either at a health facility or at home, depends on where the mother is available soon after delivery. Study research staff aim to complete birth anthropometrics within 72 hours post-delivery. Time points of data collection, biospecimen, and anthropometric assessments are provided in Table 1. For all structured interviews with the study participants, study personnel use tablet-based Research Electronic Data Capture (REDCap) applications to guide data collection customized to the visit [41, 42].

#### Demographics

Household and personal demographics captured in this study include information regarding the participant’s age, gender, sex, occupation, the language spoken in the home, religion, income, number of people this income supports, education, housing, defecation, hand washing, reproductive history and marital status. Data collection scales are adapted from the Pakistan Demographic and Health Survey (PDHS) [15].

#### Maternal empowerment and household food insecurity

An empowerment questionnaire is deployed to collect data about self-efficacy using the Generalized Self-Efficacy scale [43]. Perceived social support is measured using the Multi-dimensional Scale of Perceived Social Support (MSPSS) [44]. Perceived parental stress is measured using the Perceived Stress Scale (PSS-10) [45]. Lastly, food insecurity is assessed through the Household Food Insecurity Access Scale (HFIAS) [46]. *24-hour food recall:*

To link the microbiome to nutritional status and nutritional intake, with a focus on calories and macronutrients, an interactive semi-quantitative, 24-hour paper-based dietary recall [47] is administered by the research staff at the participants’ homes.

#### Minimum dietary diversity score for women (MDD-W)

The MDD-W is a population-level indicator for dietary diversity for women aged 15-49, based on 10 food groups. [48] The MDD-W reflects what a participant has eaten over the previous 24 hours, and participants are asked at the end of the questionnaire whether this reflects their diet over the previous 3 months. The research team will calculate the MDD-W from the dietary recalls completed at baseline, 30-34 weeks post-conception, and at 12 months.

#### Infant Feeding

WHO-developed tools are used to assess infant feeding practices [49]. At the 12-month visit, research staff administer the NutricheQ questionnaire, a tool designed for toddlers aged 1 to 3 years of age, with a focus on markers for inadequate or excessive intake and dietary imbalances [50]. Two food insecurity questions, that assess maternal and infant’s annual food insecurity are also included [46].

#### Birth history and pregnancy outcomes

Birth and labor history questions administered at the post-conception visit (24-72 hours) gather additional information on mode of delivery, gestational age, newborn anthropometrics (weight, length, head circumference), placental insufficiency, antibiotic use, among other birth characteristics.

#### Health and medicine use assessment

Morbidity assessment captures mortality, morbidity, and medication usage of mother and infant at several timepoints (Table 1).

#### Blood sample collection

Certified paramedics are trained by AKU’s faculty and senior management of Nutrition Research Laboratory (NRL) on blood collection using AKU’s Standard Operating Procedures (SOPs). Venous blood specimens are collected from participating mothers (5 mL) and infants (3 mL) and distributed into two types of vacutainers. 0.6 mL of blood transferred to an SST tube (Yellow cap BD vacutainer: BD, PL6 7BP, UK) for ferritin and c-reactive protein (CRP) analysis, while the rest will be transferred to an EDTA tube (Purple cap BD vacutainer: BD, 1 Becton Drive, Franklin Lakes, NJ 07417 USA) for HB and MCV analysis, as well as for future storage. These tubes will be transported to the field-based lab in a Coleman portable freezer maintained at 2-8°C. The extracted serum will then be flash-frozen and stored at -80°C until the point of analysis.

#### Stool sample collection

Participating mothers are provided with sterile stool containers a day before collection to provide freshly passed stool samples to field staff, while infants’ samples are collected in diapers and transferred to containers by the field team. If the stool is mixed with urine, collections are rescheduled. Collected samples are transported to the field-based lab in a Coleman portable freezer maintained at 2-8°C. At the lab, the sample is aliquoted in 4 cryovials and stored at - 80°C until further processing.

#### Anthropometry

Maternal height and weight are measured using a digital floor scale (seca 813, Seca, Hamburg, Germany) and stadiometer (seca 213); mid-upper arm circumference (MUAC) is taken using a measuring tape (seca 201). Triceps skinfold thickness (SFT) is measured using a skinfold caliper (Holtain CRYMYCH, UK). We use Seca scales for infant anthropometry, which includes measurement of weight (seca 354), length (seca 417), MUAC (seca 201), and head circumference (seca 212). All measurements are collected in duplicate, by two study personnel, using standardized procedures, as adopted from the anthropometric data collection tools used in the INTERGROWTH-21^st^ Study [51]. The average of acceptable paired measures is used in subsequent analyses.

### Patient and public involvement statement

The study participants have not been involved in the design, implementation, or analysis or dissemination plans. However, before the initiation of study, we conducted meetings with community gate keepers to orient them about the overall goal and the activities of the study.

### Data management

All tablets are synchronized daily to upload data from the REDCap data platform to a secure, web-based server hosted at the AKU campus. The tablets include built-in logic and range checks to ensure data quality. Paper-based study forms for the 24-hour food recall are checked by a study monitor for consistency and completeness. Dual entry of paper-based data is performed to reduce data entry errors. Data entry screens are developed using Visual FoxPro software (Microsoft). De-identified data is stored in a password-protected data base.

### Outcome measures

#### Primary outcome measures

To determine the weight gain, maternal weight is measured in the first and third trimesters. Gut microbiome dynamics/dysbiosis (bacterial and eukaryotic) will be assessed using 16S and 18S rDNA surveys applied to the maternal stool samples. Birth outcomes include birthweight, small for gestational age (SGA), large for gestational age (LGA), preterm births (birth before 37 week of gestation), mortality and morbidity. Infant growth and nutritional parameters include WHO z-scores for weight, length, and head circumference at birth and during infancy [52]. Beyond this we will also monitor infant morbidity, mortality, care seeking, hospitalization, antibiotic use, and feeding practices to establish association between maternal gut dysbiosis and several birth and infant outcomes.

#### Secondary outcome measures

To determine the impact of the maternal microbiome including exposure to pathogens and parasites on the development of the infant microbiome we will analyze maternal and infant microbiome composition, micronutrients in the serum (using metallomics e.g., zinc, magnesium, and selenium), and macronutrients in the stool. Maternal stool markers of intestinal mass, inflammation (Calprotectin, Lipocalin-2, Claudin-15), and gut permeability and microbiome dynamics will be used to examine the association of intestinal inflammation with microbiome’s exposure to pathogens and parasites. We will integrate maternal clinical information (i.e., morbidity, care seeking, medication use and hospitalization) and anthropometric factors with microbiome data to reveal key modulators (microbial taxa and metabolites) of dietary intake during pregnancy and the postpartum period. Household food insecurity and dietary diversity scores will be generated to link the maternal microbiome to dietary intake, with a focus on calories and macronutrients.

#### Exploratory outcomes

For the exploratory outcomes, metatranscriptomics, metabolomics and markers of inflammation will be selectively deployed on the stool samples to see the variations in dietary intake and maternal nutritional status. We will also use animal models to explore the bacterial and eukaryotic components of the microbiome. Details on outcomes measures are provided in Table 01 of supplementary material. Potential confounders are outlined under Strengths and Limitations.

### Stool analysis

Maternal and infant stool samples from all time points undergo DNA extraction at the NRL at AKU. Maternal stool RNA extraction will be completed for selected participants (100 with the highest BMI and 100 with the lowest BMI at time of enrollment) from stool collected at the two pregnancy visits. Stool samples of these participants will also be analyzed for inflammatory markers (Table 2). The stool samples and extracted DNA and RNA samples will be then batch shipped to the Hospital for Sick Children, Toronto, Canada to complete downstream analyses, including sequencing, metabolomics and biobanking.

**Table 2:** Stool assays, methods, instruments, and processing laboratories.

| Stool Analyte | Amount of stool (mg) | Assay method | Instrument | Processing Lab |
| --- | --- | --- | --- | --- |
| Calprotectin | 100 | Sandwich Immunoassay | Liaison immune analyzer, Diasorin. | NRL, AKU, Pakistan |
| Lipocalin-2 | 100 | Sandwich Immunoassay | Epoch 2 microplate reader, Biotek. | NRL, AKU, Pakistan |
| Claudin-15 | 300 | Competitive Enzyme Immunoassay | Epoch 2 microplate reader, Biotek. | NRL, AKU, Pakistan |
| RNA Extraction | 250 | Column Extraction Technology | ZymoBIOMICS™ RNA Miniprep Kit (R2001) | NRL, AKU, Pakistan |
| DNA Extraction | 200 | Column Extraction Technology | ZymoBIOMICS™ DNA Miniprep Kit (D4300) | NRL, AKU, Pakistan |
| Stool Archiving for Shipment | 2000 | - | ThermoScientific TSX-Series -80°C Ultralow temperature freezer | NRL, AKU, Pakistan |
| Stool Metabolomics* | 100 | Microbiome Metabolism Assay | LC-MS | TMIC, Alberta, Canada |
| *SCFAs, amino acids, intermediates in glycolysis and nucleotide metabolism |  |  |  |  |

### Blood analysis

Blood samples will be analyzed at AKU for hemoglobin, mean cell volume, ferritin, and CRP concentration, with additional aliquots shipped to SickKids for further analysis (Table 3). The Metabolomics Innovation Center (TMIC) will conduct metallomics analysis using the TMIC metallomics platform to investigate micronutrients (e.g., zinc, magnesium, selenium).

**Table 3:**
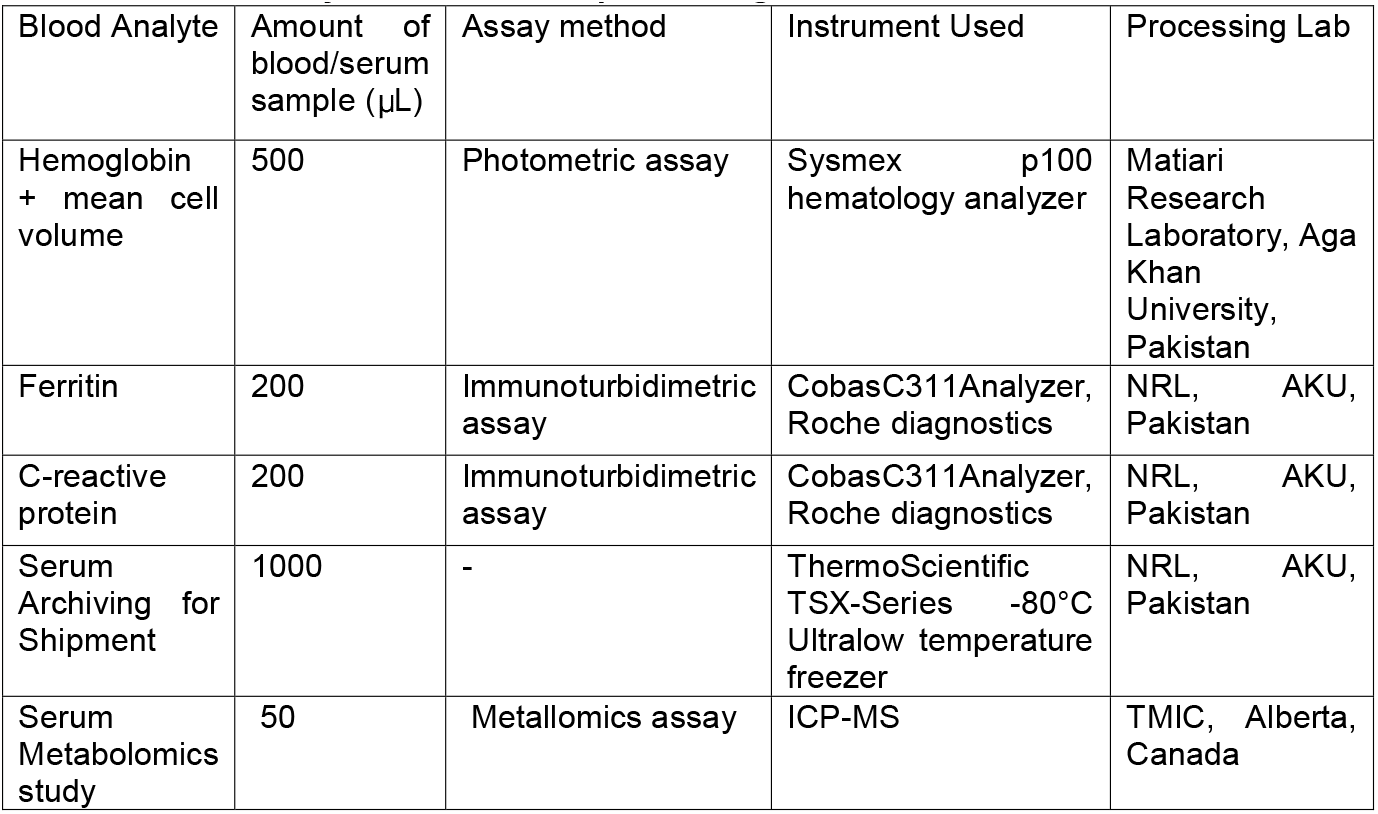
Blood assays, methods, and processing laboratories.

| Blood Analyte | Amount of blood/serum sample (μL) | Assay method | Instrument Used | Processing Lab |
| --- | --- | --- | --- | --- |
| Hemoglobin + mean cell volume | 500 | Photometric assay | Sysmex p100 hematology analyzer | Matiari Research Laboratory, Aga Khan University, Pakistan |
| Ferritin | 200 | Immunoturbidimetric assay | CobasC311Analyzer, Roche diagnostics | NRL, AKU, Pakistan |
| C-reactive protein | 200 | Immunoturbidimetric assay | CobasC311Analyzer, Roche diagnostics | NRL, AKU, Pakistan |
| Serum Archiving for Shipment | 1000 | - | ThermoScientific TSX-Series -80°C Ultralow temperature freezer | NRL, AKU, Pakistan |
| Serum Metabolomics study | 50 | Metallomics assay | ICP-MS | TMIC, Alberta, Canada |

### Profiling microbial community structure

Microbial communities will be analyzed through 16S and 18S rDNA surveys using established methods that target the V4 region of the 16S rRNA gene to capture bacterial taxa [53-55] and the V4V5 region of the 18S rRNA gene to capture eukaryotic taxa [56]. DNA library preps include error-correcting barcodes [57] for multiplexing of samples. Sequencing will be performed to generate ∼50,000 2x150bp paired end reads per sample. To define taxonomic diversity, species profiles from 16S and 18S rDNA data will be clustered to identify differences in community structure across samples. We will utilize the QIIME2 platform [58], MOTHUR [59], multivariate approaches such as Permutation Multivariate Analysis of Variance (i.e. PERMANOVA-S a method that can associate microbiome changes with outcome measures while accounting for confounders [60]. Differences between groups in microbiome community structure will be tested by analysis of similarities (ANOSIM) and co-occurrence analysis [61]. To better define bacterial pathogen burden, we apply TaqMan array card technology for the simultaneous detection of 19 common enteropathogens [62].

### Profiling microbial community function

After total RNA extraction and rRNA depletion (RiboZero Gold Kit, Illumina, San Diego, Ca, or equivalent), libraries will be constructed and Illumina-based sequencing will be performed to generate ∼30 million 2x150bp paired-end reads per sample (our rarefaction analyses have previously shown such sequencing depth is sufficient to identify the vast majority of species and enzymes present in the samples [63]). Reads will be processed for quality and contaminants using the MetaPro pipeline [64]. Reads will be assembled using SPAdes [65] and subsequently annotated with taxonomic and functional assignments. Expression will be normalized to Reads per Kilobase of transcript per million mapped reads (RPKM). Annotations will be mapped onto biochemical pathways and complexes such as those defined by the Kyoto Encyclopedia of Genes and Genomes [66]. The output of these analyses will be readouts of microbial gene expression detailing biochemical activities as well as the taxa responsible.

### Statistical Analysis

Normally distributed continuous data will be shown as a mean and standard deviation, and median and Interquartile range (IQR) will be calculated for non-normally distributed data. Categorical data will be presented using proportions.

For the primary outcomes, maternal gut bacteria and eukaryotic profiles will be used to calculate Bray-Curtis dissimilarity metrics between individual samples which is leveraged in principal co-ordinate analyses to determine the extent samples collected at the first or third trimester, exhibiting similar gestational weight gains, co-cluster. Permutational multivariate analysis of variance (PERMANOVA) tests will assess the degree of overlap between samples exhibiting low gestational weight gain versus samples exhibiting high gestational weight gain. Next, we will attempt to correlate changes in the alpha diversity (as measured by the Shannon and Simpson indices) of the gut microbiome samples between the first and third trimester, with gestational weight gain. To examine the influence of individual taxa on gestational weight gain, we will perform bivariate analyses (Pearson, Spearman). The Benjamini-Hochberg procedure will be applied to correct p-values while controlling for false-discovery rates.

To complement these analyses, we will also undertake an integrative modeling strategy based on the Similarity Network Fusion framework [67] to analyze the contribution of each variable (clinical, microbiome, and gender-related, see Table 1 of supplementary material) on gestational weight gain. This allows the integration of all available datasets to uncover their global substructures that can be associated with gestational weight gain. In an alternative approach, we will also employ Random Forests to identify combinations of variables that correlate with gestational weight gain.

For the rest of the outcomes, followed by general linear models i.e., PERMANOVA and the Bray-Curtis dissimilarity metric. DESeq2, a method for differential analysis of count data, will be applied to investigate both associations of specific taxa with the clinical variables, together with the strength of those associations. These analyses reveal which clinical variables (including exposure to pathogens and parasites) correlate with the maternal microbiome from a taxonomic perspective. Additionally, microbiome structural and functional profiles will be generated from the difference in 1) taxonomic abundances; 2) gene expression; and 3) metabolite concentrations, between the first and third trimesters. Profile differences will be utilized in the PERMANOVA and DESeq2 approaches as described above to identify associations between clinical variables and changes in microbiome structure and function. Throughout these analyses we will include potential confounders such as medication use, impact of flooding and time of sample collection as outlined in strengths and limitations, as covariates where appropriate. Where consented, patient DNA data offers additional opportunities to integrate host genetics into these analyses (see supplementary material).

## Discussion

Nutritional status during pregnancy plays an important role in maternal health and birth outcomes [68, 69]. Maternal undernutrition during pregnancy can lead to fetal growth restriction, which increases the risk of neonatal deaths and childhood stunting by 2 years of age [2]. In Pakistan, large scale surveys and cohort studies have suggested multifaceted undernutrition and adverse pregnancy and birth outcomes [13-15]. Data from a cohort of young women (15-24 years) living in rural Pakistan revealed that more than 90 percent lives with minimum one micronutrient deficiency [36] and nearly 40% were underweight [34].. Dietary intake was limited to fewer types of foods, mainly staples [35]

The gut microbiome can have a profound influence on host’s nutritional status, yet few studies of the dynamics between nutritional status and the gut microbiome during pregnancy have been conducted. Further, a few studies focusing on child undernutrition have revealed a key role for gut microbiota [16-22]. In particular, dysbiosis, or the loss of diversity/beneficial microbes and gain of pathobionts, has emerged as a major factor in the development of undernutrition. To date most studies of the gut microbiome have focused on bacterial components, typically neglecting the contribution of eukaryotic microbiota. Despite the fact that many such eukaryotes include parasites, such as *Giardia, Cryptosporidium*, and *Entamoeba*, each representing a significant burden on global healthcare with considerable implications for gut health [70-73]. Interestingly, not all parasitic infections cause disease; instead, many infections remain asymptomatic with disease emerging as a consequence of interactions between the eukaryotic and bacterial microbiome and the host immune system [74, 75]. With the emergence of new marker gene technology, based on the 18S/5S/28S locus, there is now the opportunity to profile eukaryotic communities and examine their impact within the context of the gut microbiome.

Thus, this study will inform the relationships between host nutritional status and microbiome dynamics during pregnancy, and how they contribute to gestational weight gain during pregnancy, in addition to several other pregnancy and birth-related outcomes. Understanding the role of the microbiome on maternal health and birth outcomes, as well as the influence of enteric eukaryotic microbes, such as parasites, on the bacterial microbiome and host nutrition offers great potential in the identification of modifiable factors to improve health and nutrition outcomes.

To help establish causal relationships between microbiome dynamics, pathogen exposure, and nutritional status during pregnancy and to examine whether manipulation of the microbiome can improve nutritional status, future work is expected to leverage stool samples collected here, in fecal microbiome transplant studies using animal models.

## Strengths and limitations

The study targets the high fertility age group (17-24) with almost half cohort consist of low BMI mothers, potentially with an additional risk of adverse pregnancy outcomes, providing an opportunity to comprehend the systematic understanding of the role of microbiota in several pregnancy, birth, and infant outcomes. Study investigates both prokaryotic and eukaryotic dynamics of the gut microbiome for in-depth mechanistic insights in a highly malnourished population where contextual evidence is rare. The longitudinal design and data collection on a range of exposure indicators and biochemical analysis would enable the analysis to evaluate the association of gut dynamics with several physiological and environmental factors. The study follows the STROBE guidelines [76]; however, we expect controlling for all biases and confounding variables may not be possible. For example, verbal data is collected through interviews, while some follow ups collect three to six months recall data on morbidity, medication use and care seeking which may impact the reliability of reporting mothers. However, to increase accuracy of reporting, participants are encouraged to keep the record of medicines i.e., prescriptions and reports. We have developed a pictorial list of medicine to improve mothers’ recall. Of particular note, the study population experienced an unprecedent flooding event in 2022 which disrupted daily life and dietary patterns. Further, the flooding event is also expected to have increased exposure of the study population to additional pathogens, with corresponding impacts on microbiome dynamics. We anticipate performing a subgroup analysis to investigate if exposure to flooding had a significant impact on microbial dynamics. Due to restricted hours of operation relative to the passing of stool by infants, we expect that there may be heterogeneity in the amount of time between passing of stool and collection by the field team. At the same time, during sample collection we ensure that the specimen temperature is maintained until it reaches the field-base laboratory. During analysis, we may find that we identify no significant differences between microbial diversity or composition in relation to our primary or secondary outcomes. Such findings would elevate the importance of the metatranscriptomic analyses to deliver more mechanistic investigations. It is possible that RNA quality and yields from stool is poor. In such events we will revert to performing whole microbiome DNA (which is more stable than RNA) sequencing (metagenomics) which also has the capacity to deliver functional insights. Finally, we acknowledge that by focusing on young women, 17-24 years of age, that findings may not be generalizable to younger or older demographics. At the same time, studies of the microbiome within this age group are lacking and hence this study is aimed at directly addressing this knowledge gap to deliver a wealth of information to better serve the health care needs of this important demographic.

## Data Availability

All data produced in the present study are available upon reasonable request to the authors

## Ethics

This study was approved by the national bioethics committee (NBC) in Pakistan (NBC Ref: No.4-87/NBC-700/21/820), the institutional ethics review committee (ERC) at AKU (ERC No.2021-6085-17561) and the research ethics board (REB) at the Hospital for Sick Children (SickKids; REB number: 1000076773).

## Dissemination

The study is registered with ClinicalTrials.gov Identifier: NCT05108675. Results will be published in open access peer-reviewed journals.

## Authors’ contributions

Dr. Zulfiqar Ali Bhutta (PI) and JP envisioned the study and got funding. CS, JBB, JH, RB, SS, SBS, YW and JP drafted the protocol. KB and JI helped finalize laboratory work for Pakistan based cohort. AR assisted with data analysis plan. YW, JBB, CS developed questionnaires and field SOPs. YW produced initial draft of this manuscript with inputs from team mentioned above. All the listed authors reviewed and approved final draft for publication.

## Acknowledgements

We are delighted to acknowledge the valued contribution of study field team including Ms. Azra Sheikh, Mr. Abdullah Memon and Mansoor Ali Abro for translating and implementing the data collection questionnaires in local language. We are highly indebted to Mr. Imran Memon and Amjad Hussain for their invaluable assistance with the development of data collection applications. Our special thanks to Mr. Aadil Qureshi and Rafique Soomro for their unwavering onsite admin and logistics support to run the research operations. We also greatly value the contribution of Mr. Shahneel Hussain for ensuring quality standards and logistics for specimen collection. We appreciate the key contribution of grant and administration team, data management unit, and several other departments at AKU for successfully initiating this study in the field. Last but not least, we extend our gratitude to the families who voluntarily agreed to participate in this study.

## Abbreviations

ANOSIM: Analysis of Similarities
BMI: Body Mass Index
CRP: C-reactive protein
DNA: Deoxyribonucleic acid
EDTA: Ethylenediamine tetraacetic acid
EED: Environmental Enteric Dysfunction
ERC: Ethics Review Committee
HFIAS: Household Food Insecurity Access Scale
IV: Infant Visit
LBW: Low Birth Weight
LMICs: Low and Middle-income Countries
MAL-ED: Malnutrition and Enteric Disease
MaPPS: Matiari Empowerment and Preconception Supplementation
MCV: Mean Corpuscular Volume
MDD-W: Minimum dietary diversity score for women
MSPSS: Multi-Dimensional Scale of Perceived Social Support
MUAC: Mid Upper Arm Circumference
MV: Maternal visit
NBC: National Bioethics Committee
NRL: Nutrition Research Laboratory
PDHS: Pakistan Demographic and Health Survey
PERMANOVA: Permutational Multivariate Analysis of Variance
QIIME: Quantitative Insights into Microbial Ecology
RPKM: Reads per Kilobase of transcript per million mapped reads
REB: Research Ethics Board
REDCap: Redcap Research Electronic Data Capture
RPKM s: Reads Per Kilobase of Transcript Per Million Mapped
RNA: Ribonucleic acid
SCFA: Short-chain fatty acids
SFT: Triceps Skinfold Thickness
SOPs: Standard Operating Procedures
SST: Serum Separator Tube
STROBE: Strengthening the reporting of observational studies in epidemiology
TMIC: The Metabolomics Innovation Center
WHO: World Health Organization

