## Supplementary Material for "Elucidating the Dynamics and Impact of the Gut Microbiome on Maternal Nutritional Status During Pregnancy in Rural Pakistan: Study Protocol for a Prospective, Longitudinal Observational Study"

**Future Genetic Testing:**

Although not funded as part of this study, given the lack of data to link genomics with gut we envision that the availability of DNA samples from the mothers and children enrolled in this study provides an unparalleled opportunity to apply whole genome sequencing or targeted resequencing methodology, to examine how the interactions between host genetic factors and the gut microbiome contribute to host nutritional status and birth outcomes. Such studies would form the basis of future exhaustive investigations.

**Table 1: Objectives and outcome measures**

| Objectives | Outcome measures | Comments |
|---|---|---|
| Primary | | |
| To assess if alterations of the microbiota in the maternal gut (dysbiosis) are associated with maternal gestational weight gain. | - Gestational weight gain<br>Microbiome dynamics:<br>- Bacterial and eukaryotic components of the microbiome. Using 16S and 18S rDNA surveys on maternal stool samples | Weight gain is a key predictor of birth outcomes. |
| To determine the association between maternal microbiome dysbiosis during pregnancy and birth outcomes, infant growth, nutritional status, and health status in the first year of life. | Birth outcomes:<br>- Birth weight<br>- Preterm birth<br>- small-for- and large-for-gestational age<br>- Head circumference at birth<br>- Mortality<br>- Morbidity<br>Infant outcomes:<br>Growth and nutritional status parameters:<br>- WHO z-scores for weight length, head circumference, and MUAC during the first year.<br>- Infant morbidity<br>- Infant mortality<br>- Care seeking<br>- Antibiotic use<br>- Hospitalization<br>- Feeding practices<br>Microbiome dynamics:<br>- Bacterial and eukaryotic components of the microbiome.<br>- Using 16S and 18S rDNA surveys on maternal stool samples | This will allow for an examination of the association between the maternal microbiome and birth and infant outcomes. |
| Secondary | | |

| Objectives | Outcome measures | Comments |
| --- | --- | --- |
| To link the maternal microbiome to dietary intake, with a focus on calories and macronutrients. | - Dietary diversity scores<br>- Household food insecurity | Quantitative measures of nutritional status will provide parameters to better inform microbiome analyses. |
| To integrate maternal anthropometric factors, morbidity, and mortality with microbiome data to reveal key modulators (microbial taxa and metabolites) of dietary intake during pregnancy and the postpartum period. | - Maternal morbidity<br>- Complications<br>- Care seeking<br>- Hospitalization<br>- Medication use<br>- Maternal BMI | Integration with clinical information will allow for the analysis of microbiome data in the context of patient nutritional status and health outcomes. |
| To determine the impact of the maternal microbiome during pregnancy, including the exposure to pathogens and parasites, on the development of the infant microbiome. | - Maternal and infant microbiome composition<br>- Micronutrients in the serum (e.g., zinc, magnesium, and selenium)<br>- Macronutrients in stool | These analyses will inform on the role of the maternal microbiome during pregnancy to impact the developing child's microbiome. Our hypothesis is that exposure to pathogens and/or parasites during pregnancy will negatively impact the development of the child's microbiome with downstream consequences for nutrient uptake. |
| To investigate the maternal microbiome's exposure to pathogens and parasites, and the association with intestinal inflammation. | Maternal stool markers of intestinal mass, inflammation, and gut permeability:<br>- Calprotectin<br>- Lipocalin-2<br>- Claudin-15<br>Microbiome dynamics:<br>- Bacterial and eukaryotic components of the microbiome<br>- Using 16S and 18S rDNA surveys on maternal stool samples | These markers will reflect gut health and pathogen challenges. |
| Exploratory | | |

| Objectives | Outcome measures | Comments |
| --- | --- | --- |
| To explore the role of the maternal gut microbiome during pregnancy and to identify gut community dynamics in pregnant women and how this impacts differences between dietary intake and nutritional status. | - Metatranscriptomics<br>- Metabolomics and markers of inflammation will be selectively deployed on the stool samples | The output of these analyses are concentrations of metabolites that are expected to correlate with pathway expression data; linked to readouts of microbial gene expression detailing biochemical activity and the taxa responsible. |
| To investigate socio-economic factors; including gender, poverty, exclusion, and empowerment, and their influence on the health of a mother's microbiome (assessed by alpha and/or beta diversity, and absence of pathogens). | Gender-related variable:<br>- Perceived maternal self-efficacy<br>- Perceived social support, decision making<br>- Perceived stress<br>- Wealth quantiles<br>- Household food insecurity<br>- Education<br>- Household density | The use of previously validated questionnaires within this population, will allow for comparison between the Pakistan and Toronto cohorts, to account for gender related variables that impact maternal and infant gut health. |
| To explore the role of the human microbiota on nutritional status by performing fecal microbiota transplants in germ-free mice and sterile piglets. | - Pregnancy rates<br>- Birth outcomes<br>- Microbiome dynamics (bacterial and eukaryotic components of the microbiome)<br>- Health outcomes (morbidity and mortality)<br>- Host nutritional status<br>- Inflammation<br>- Barrier integrity | Exploiting animal models will allow us to define causal interactions between diet, microbiome, pathogen exposure and nutritional status during pregnancy. Through recapitulating patient phenotypes with fecal microbiome transplants, our animal studies will set the stage for developing new therapeutic strategies that promote gut health. |
